# The Uncertain COVID-19 Spread Pattern in India: A Statistical Analysis of the Current Situation

**DOI:** 10.1101/2020.08.30.20184598

**Authors:** Hemanta K. Baruah

**Affiliations:** Department of Mathematics, The Assam Royal Global University, Guwahati, Assam, India

**Keywords:** Pandemic, infectious disease, regression analysis

## Abstract

There are standard techniques of forecasting the spread of pandemics. Uncertainty however is always associated with such forecasts. In this article, we are going to discuss the uncertain situation currently prevailing in the COVID-19 spread in India. For statistical analysis, we have considered the total number of cases for 60 consecutive days, from June 23 to August 21. We have seen that instead of taking data of all 60 days together, a better picture of uncertainty can be observed if we consider the data separately in three equal parts from June 23 to July 12, from July 13 to August 1, and from August 2 to August 21. For that we would first need to ascertain that the current spread pattern in India is almost exponential. Thereafter we shall show that the data regarding the total number of cases in India are not really behaving in an expected way, making forecasting the time to peak very difficult. We have found that the pandemic would perhaps change its pattern of growth from nearly exponential to nearly logarithmic, which we have earlier observed in the case of Italy, in less than 78 days starting from August 2.

**AMS Mathematics Subject Classification (2010):** 97K70

## 1. Introduction

The epidemiological mathematical models commonly used to forecast the spread of a pandemic are the Susceptible-Infectious-Recovered (SIR) model [1, 2, 3] and its modifications and generalizations such as the Susceptible-Exposed-Infectious-Recovered (SEIR) model, the Susceptible-Infectious-Recovered-Dead (SIRD) model, the Susceptible-Infectious-Susceptible (SIS) model etc. Such models are applied to forecast the total number of infected cases and the duration that the pandemic may be expected to continue to grow. These models are used to estimate the basic reproduction number known as *R*_0_with threshold properties. It is the mean number of infections caused by one single individual in a susceptible population. If this number is found to be greater than1, it would mean that the population would remain infected permanently, and if it is found to be less than or equal to1, it would mean that the disease would extinct in course of time. In the classical SIR model, it is assumed that the duration spent by an individual in the infectious state follows the exponential probability distribution. The basic reproduction number was defined as

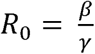

where *β* ^−1^is the typical time between contacts and *γ*^−1^is the typical time until recovery. It may be noted that these models are mathematically valid when the population is assumed to be infinite.

Therefore although in a small region this model can be expected to work well, in a very large region application of this model may perhaps be unrealistic. In a large region, medical facilities may not be uniformly available for everyone. Economic disparities would certainly affect susceptibility to the disease. This may lead to an imperfect value of the basic reproduction number. Therefore the validity of such models with an assumption that the population is infinite is questionable when the disease is COVID-19 in particular for which medicine is yet to be made available to everyone.

The SIS model is used to study infectious diseases that do not last very long, and it assumes that such infections do not lead to immunity after recovery. The SIRD model typically assumes that the recovered cases are immune to the disease. In the SEIR model, it is assumed that there is a period of incubation within which infected individuals are not infectious. Individuals that are infected but not infectious are considered as exposed in this model.

As can be seen, COVID-19 has lasted for eight months now, and it is uncertain as yet how long it would continue the world over. Therefore the SIS model is perhaps unfit to forecast about the COVID-19 spread because in this model it is assumed that the disease would not last very long. It has actually been reported that some people who had recovered from the disease have been infected again, although the number may be quite low. As such, a presumption that the recovered cases are immune to the disease is debatable. Therefore the SIRD model may not perhaps lead to good forecasts. Without statistical tests, rejecting the hypothesis that within the incubation period infected individuals are not infectious may not be proper. But in the COVID-19 spread related works in which the SEIR model was used it was apparently presumed that within the incubation period the infected individuals are not carriers of the disease. We therefore cannot be sure that application of the SEIR model is perfect in the COVID-19 spread matters.

Indeed, from the graphical representation of the total number of COVID-19 cases in the USA as shown by the Worldometers.info [4] data, it is apparent that unlike in the cases of any other country, in the USA the shape of the curve has not been showing a pattern as presumed in the epidemiological models. In the USA, the curve has not been showing a smooth increase. Therefore application of such models in the case of the USA may perhaps need modification of the models.

There were attempts to study the spread pattern using the auto-regressive integrated moving average (ARIMA) method also. Indeed, in the initial stage, forecasting with the help of ARIMA did lead to very good short term forecasts. For example, Poonia and Azad [5] and Azad and Poonia [6] have studied forecasting of the disease in India in two phases using the ARIMA method. Their forecasts were very close to the values observed later. These two works were done at the very initial stage of the outbreak of the disease. However, we have observed that for long term forecasting, this method in this kind of a pandemic situation may not be very suitable.

We would now like to cite some more works done by various researchers with reference to COVID-19 spread in India. Pai *et. al*. [7] have studied the COVID spread matters in India using the SEIR model. Gupta *et. al* [8] also have studied the situation in India using the SEIR model as well as regression analysis. Ranjan [9] studied about predictions in India using epidemiological models. Acharya and Powal [10] have done an ecological study of the Indian situation with reference to a vulnerability index. Their studies were however based on data till the middle of June. They have observed similarities between vulnerability and the concentration of the COVID-19 cases at the state level of India. All these works on the Indian situation were done in the beginning of the spread of the disease.

It can be observed from the diagrammatic representation of the data of the total number of cases of COVID-19 in India is currently following an almost exponential pattern. The pattern that looks exponential can actually be almost exponential, and *not* exactly exponential. In the population dynamics models an S-type of spread curve is used in which the growth is slowest in the beginning and at the end of a given duration. In the case of the COVID-19 spread in every individual region, the growth at the start was nonlinear. In the regions in which the spread has reached the final stage of retardation, it has been found to be slowest once again. Between these two stages, there are two stages, one with nearly exponential growth and the next is with nearly logarithmic growth.

The spread pattern in India is still in the nearly exponential stage. In India the third stage is yet to come. In this article, we are going to show that instead of considering an epidemiological model of the types discussed above, we may proceed to study the spread of a pandemic part by part with reference to time. To know whether the spread is about to retard from an exponential pattern to a logarithmic pattern, we first need to study whether the growth during the nearly exponential period is about to change.

## 2. Methodology

It can be observed from the diagrammatic representation of the daily data of the total number of cases of COVID-19 in India that the total number of cases is following an almost exponential pattern. Indeed, the spread pattern of a pandemic in any region can never be exactly exponential, for that would mean that the pandemic would never come to a stop. Therefore, the pattern that looks exponential may actually be almost exponential, and not exactly exponential.

For a function *g*(*t*), exponential in *t*, let us write

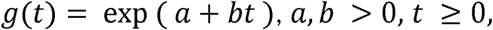

where *a* and *b* are constants so that

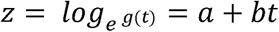

is linear in *t*. If we want to use this model in the exponential stage of the spread of a pandemic, we would need to take some chosen value of *t* and from that *t* as base we can proceed to find the value of *b*. Accordingly, the value of the constant *a* would be available to us already and we would have to find an estimate of the parameter *b*. When we would observe that the estimated values of *b* are very nearly constant, we would be able to say that the pattern is approximately exponential.

It is apparent from the graph published by Worldometers.info [4] that in India the spread pattern is still approximately exponential. It was seen that the function

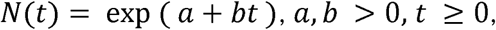

fits the data of spread in India approximately [11, 12, 13]. Let us write *z*(*t*)= *log*_*e*_ *N*(*t*).

To estimate the value of the parameter *b* at some point of time we would need data about the total number of cases for a few days prior to that. If the values of Δ*z*(*t*), the first order differences of *z*(*t*) are seen to be nearly constant, then we can say that the pattern is nearly exponential.

It was observed in [11, 12, 13] that in the case of India, the values of Δ*z*(*t*) have been following a reducing trend with some irregularities in between which is inherent in the case of a time series of this type of a pandemic. The average value of Δ*z*(*t*) in India [11] during the 14 days from May 11 to May 24 was 0.051716. It was seen further [11] that during the 7 days from May 25 to May 31, the average value of Δ*z*(*t*) was 0.045584. Therefore the reducing trend of the values of Δ*z*(*t*) was apparent in those 21 days. The average value of Δ*z*(*t*) in India [12] from June 1 to June 7, 2020, was 0.04063. From June 8 to June 10, the average [13] came down to 0.03635. The average values of Δ*z*(*t*) were coming down slowly and steadily as could be seen.

The question is: how far would Δ*z*(*t*) continue to decrease before the spread pattern becomes nearly logarithmic. It cannot continue to decrease to zero because if that is the case, then the total number of cases would suddenly have to become a constant. The S-type curve for the epidemiological models does not say that. Therefore the change from approximately exponential to approximately logarithmic pattern must take place much before Δ*z*(*t*) becomes zero. For Italy, it was seen that [13] the change from the exponential to the logarithmic pattern took place before April 30. We are now interested to see whether India is about to be in that transition from exponential to logarithmic stage examining the data from June 23 to August 21, 2020.

## 3. Analysis and discussions

We now proceed to study the current Indian COVID-19 spread situation. We have collected the data from Worldometers.info [4]. The data considered for analysis were for 60 consecutive days from June 23 to August 21, 2020. So as to observe the trend of Δ*z*(*t*), we divided the data into three equal parts each of 20 days, from June 23 to July 12, from July 13 to August 1, and from August 2 to August 21. In Table-1, we have shown the data for the first 20 days, in Table-2 for the next 20 days and in Table-3 for the last twenty days. In each of these three tables *N*(*t*) represents the total number of COVID-19 cases till the particular date shown in the first column of every table, *z*(*t*) represents the natural logarithm of *N*(*t*), and Δ*z*(*t*) represents the first order differences of *z*(*t*). We have divided the data into three distinct parts each of duration 20 days because we were interested to see whether in every of these three durations Δ*z*(*t*) had actually been decreasing, and if so what sort of a decreasing trend was Δ*z*(*t*) following. The manner in which the pandemic has been spreading in India, we presumed that data of 60 days should be sufficient to see whether it is time for the growth curve to transform into a nearly logarithmic shape. Indeed, just the first order differences of the total number of cases *N*(*t*) may not immediately give a correct picture. The cumulative total has been growing nearly exponentially since the month of May [11]. Therefore Δ*N*(*t*) does not immediately reflect a clear picture.

**Table-1:**
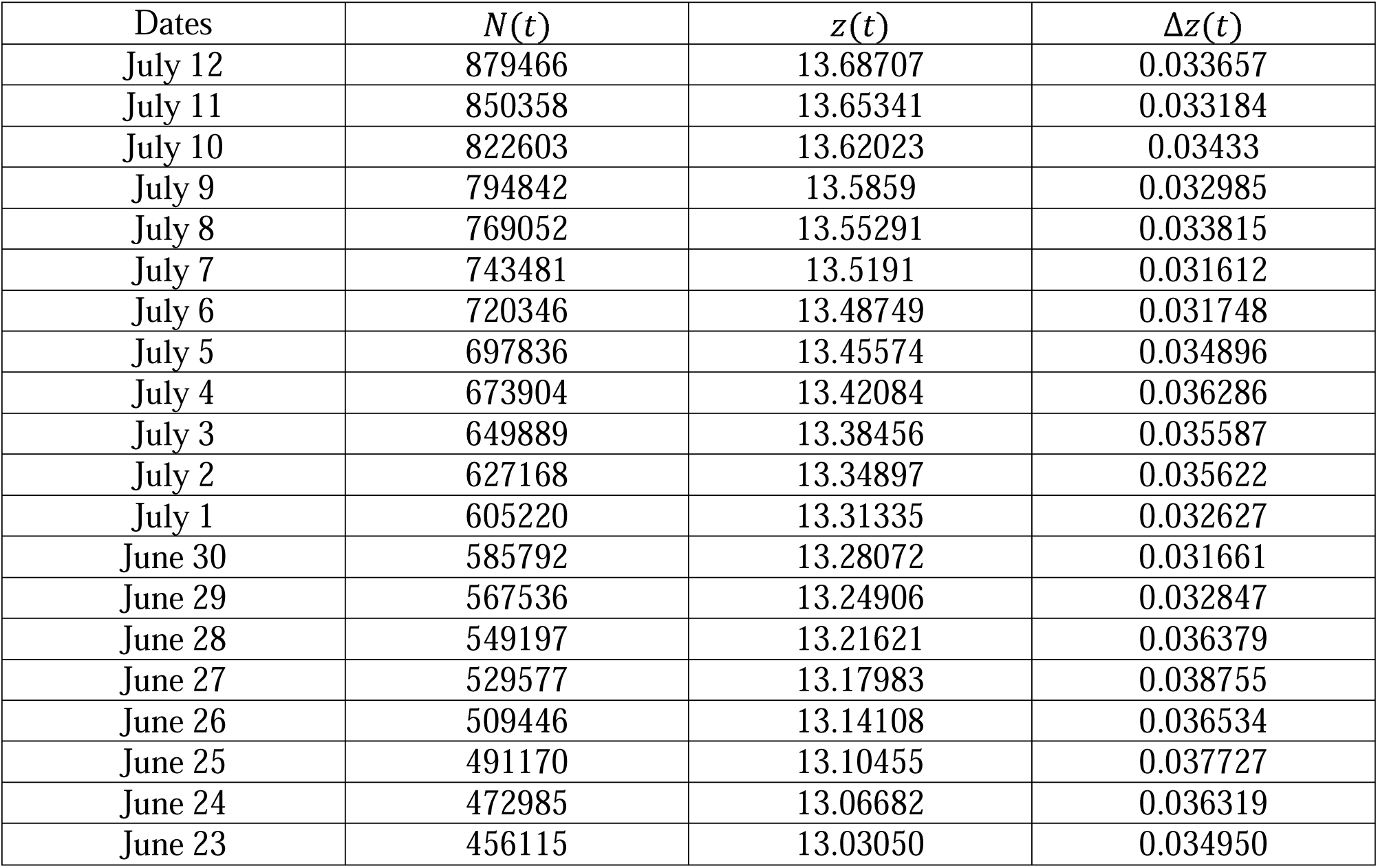
Values of Δ*z*(*t*) from June 23 to July 12

| Dates | $N(t)$ | $z(t)$ | $\Delta z(t)$ |
| --- | --- | --- | --- |
| July 12 | 879466 | 13.68707 | 0.033657 |
| July 11 | 850358 | 13.65341 | 0.033184 |
| July 10 | 822603 | 13.62023 | 0.03433 |
| July 9 | 794842 | 13.5859 | 0.032985 |
| July 8 | 769052 | 13.55291 | 0.033815 |
| July 7 | 743481 | 13.5191 | 0.031612 |
| July 6 | 720346 | 13.48749 | 0.031748 |
| July 5 | 697836 | 13.45574 | 0.034896 |
| July 4 | 673904 | 13.42084 | 0.036286 |
| July 3 | 649889 | 13.38456 | 0.035587 |
| July 2 | 627168 | 13.34897 | 0.035622 |
| July 1 | 605220 | 13.31335 | 0.032627 |
| June 30 | 585792 | 13.28072 | 0.031661 |
| June 29 | 567536 | 13.24906 | 0.032847 |
| June 28 | 549197 | 13.21621 | 0.036379 |
| June 27 | 529577 | 13.17983 | 0.038755 |
| June 26 | 509446 | 13.14108 | 0.036534 |
| June 25 | 491170 | 13.10455 | 0.037727 |
| June 24 | 472985 | 13.06682 | 0.036319 |
| June 23 | 456115 | 13.03050 | 0.034950 |

**Table-2:** Values of Δ*z*(*t*) from July 13 to August 1

| Dates | $N(t)$ | $z(t)$ | $\Delta z(t)$ |
| --- | --- | --- | --- |
| August 1 | 1751919 | 14.37622 | 0.031818 |
| July 31 | 1697054 | 14.3444 | 0.034594 |
| July 30 | 1639350 | 14.30981 | 0.034104 |
| July 29 | 1584384 | 14.27571 | 0.033534 |
| July 28 | 1532135 | 14.24217 | 0.03293 |
| July 27 | 1482503 | 14.20924 | 0.031857 |
| July 26 | 1436019 | 14.17739 | 0.035818 |
| July 25 | 1385494 | 14.14157 | 0.035612 |
| July 24 | 1337022 | 14.10596 | 0.037253 |
| July 23 | 1288130 | 14.0687 | 0.038335 |
| July 22 | 1239684 | 14.03037 | 0.037476 |
| July 21 | 1194085 | 13.99289 | 0.033352 |
| July 20 | 1154917 | 13.95954 | 0.031676 |
| July 19 | 1118907 | 13.92786 | 0.037371 |
| July 18 | 1077864 | 13.89049 | 0.035321 |
| July 17 | 1040457 | 13.85517 | 0.034039 |
| July 16 | 1005637 | 13.82113 | 0.035906 |
| July 15 | 970169 | 13.78523 | 0.034267 |
| July 14 | 937487 | 13.75096 | 0.03235 |
| July 13 | 907645 | 13.71861 | 0.031539 |

**Table-3:** Values of Δ*z*(*t*) from August 2 to August 21

| Dates | $N(t)$ | $z(t)$ | $\Delta z(t)$ |
| --- | --- | --- | --- |
| August 21 | 2973368 | 14.90521 | 0.023493 |
| August 20 | 2904329 | 14.88171 | 0.023871 |
| August 19 | 2835822 | 14.85784 | 0.024703 |
| August 18 | 2766626 | 14.83314 | 0.023783 |
| August 17 | 2701604 | 14.80936 | 0.020299 |
| August 16 | 2647316 | 14.78906 | 0.022194 |
| August 15 | 2589208 | 14.76686 | 0.025023 |
| August 14 | 2525222 | 14.74184 | 0.026325 |
| August 13 | 2459613 | 14.71551 | 0.026424 |
| August 12 | 2395471 | 14.68909 | 0.028396 |
| August 11 | 2328405 | 14.66069 | 0.026659 |
| August 10 | 2267153 | 14.63404 | 0.023662 |
| August 9 | 2214137 | 14.61037 | 0.028456 |
| August 8 | 2152020 | 14.58192 | 0.030744 |
| August 7 | 2086864 | 14.55117 | 0.029891 |
| August 6 | 2025409 | 14.52128 | 0.031176 |
| August 5 | 1963239 | 14.49011 | 0.029267 |
| August 4 | 1906613 | 14.46084 | 0.027265 |
| August 3 | 1855331 | 14.43357 | 0.027668 |
| August 2 | 1804702 | 14.40591 | 0.029684 |

In Table-4, we have shown the calculated values of the first order differences Δ*z*(*t*) for days from 1 to 20 in the three periods. This we have done so that we can visually compare the values to see if there actually is a decreasing trend in the values ofΔ*z*(*t*).

**Table-4:** Summarized form of the Tables-1, 2 and 3

| Days | Values of $\Delta z(t)$ from June 23 to July 12 | Values of $\Delta z(t)$ from July 13 to August 1 | Values of $\Delta z(t)$ from August 2 to August 21 |
| --- | --- | --- | --- |
| 20 | 0.033657 | 0.031818 | 0.023493 |
| 19 | 0.033184 | 0.034594 | 0.023871 |
| 18 | 0.03433 | 0.034104 | 0.024703 |
| 17 | 0.032985 | 0.033534 | 0.023783 |
| 16 | 0.033815 | 0.03293 | 0.020299 |
| 15 | 0.031612 | 0.031857 | 0.022194 |
| 14 | 0.031748 | 0.035818 | 0.025023 |
| 13 | 0.034896 | 0.035612 | 0.026325 |
| 12 | 0.036286 | 0.037253 | 0.026424 |
| 11 | 0.035587 | 0.038335 | 0.028396 |
| 10 | 0.035622 | 0.037476 | 0.026659 |
| 9 | 0.032627 | 0.033352 | 0.023662 |
| 8 | 0.031661 | 0.031676 | 0.028456 |
| 7 | 0.032847 | 0.037371 | 0.030744 |
| 6 | 0.036379 | 0.035321 | 0.029891 |
| 5 | 0.038755 | 0.034039 | 0.031176 |
| 4 | 0.036534 | 0.035906 | 0.029267 |
| 3 | 0.037727 | 0.034267 | 0.027265 |
| 2 | 0.036319 | 0.03235 | 0.027668 |
| 1 | 0.034950 | 0.031539 | 0.029684 |

From June 23 to July 12, the average value of Δ*z*(*t*) was 0.034576, from July 13 to August 1 it was 0.034458, and from August 2 to August 21 it was 0.026449. Period-wise the average looks to have come down observably, particularly in the last 20 days of the period concerned.

We have found that Δ*z*(*t*) during these three periods follows the following three linear equations established using the method of least squares. From June 23 to July 12,

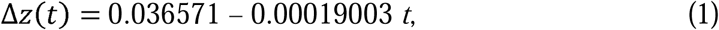

from July 13 to August 1,

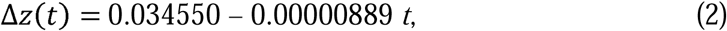

and from August 2 to August 21,

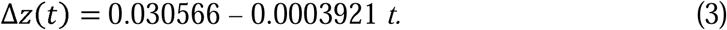

It is apparent that in the three regression equations of Δ*z*(*t*) on *t* above, the slopes are indeed negative, but the retardations are very slow. We have done the statistical tests of significance of the null hypothesis *H*_0_: *ρ*= 0 against the two sided alternative hypothesis : *H*_1_ *ρ*≠ 0 where is the population correlation coefficient between the variables Δ*z*(*t*) and *t* for the three periods mentioned with reference to equations (1), (2) and (3).

In Fig. 1, we have shown a diagrammatic comparison of the values of Δ*z*(*t*) calculated using equation (3) and the corresponding observed values shown as a broken line of Δ*z*(*t*) from Table-3, for the period from August 2 to August 21.

**Fig. 1:**
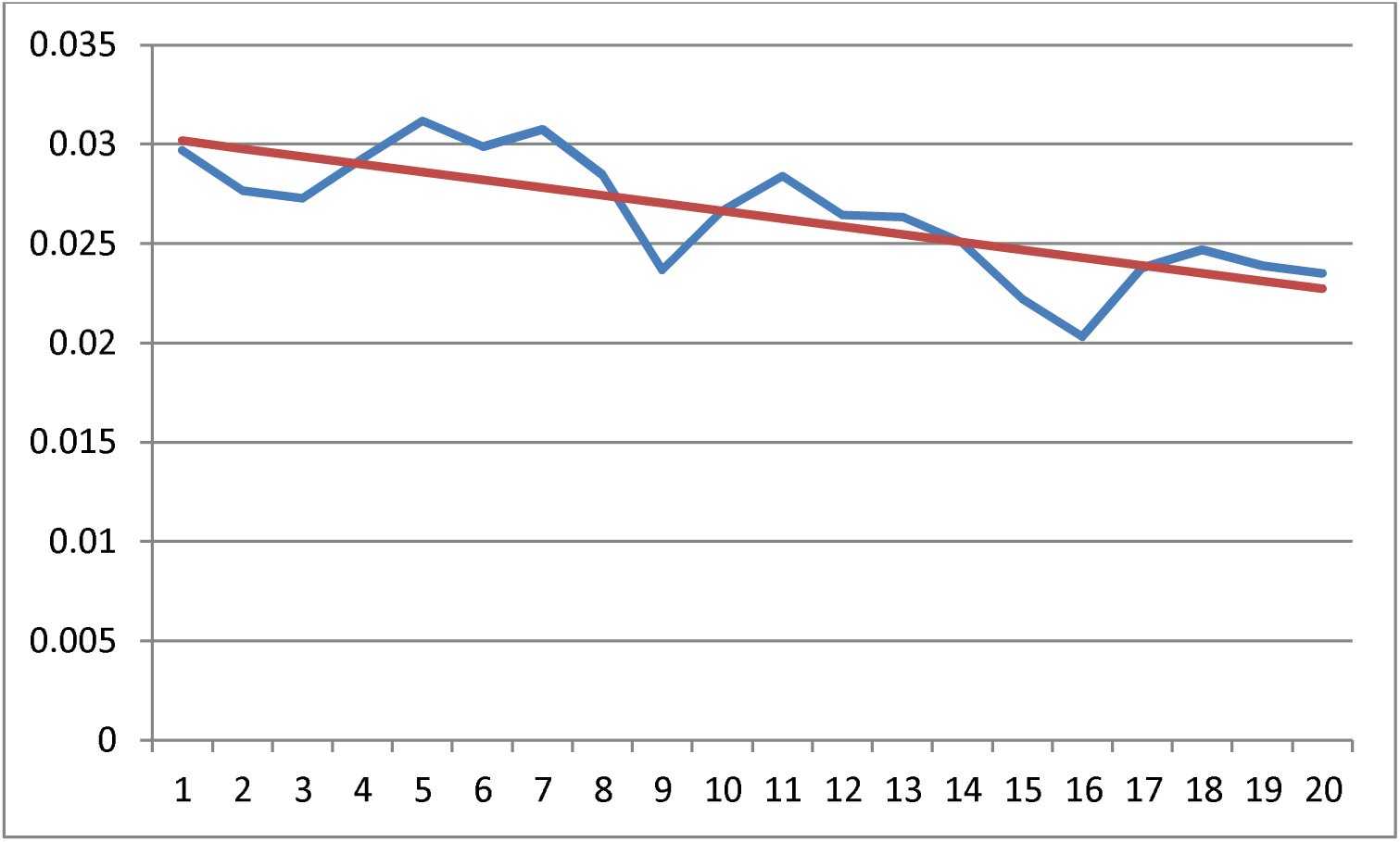
Comparison of the expected and the observed values of Δ*z*(*t*) from August 2 to August 21

It has been found that for the period from June 23 to July 12, *r*^2^= 0.2947194, for the period from July 13 to August 1, *r*^2^ = 0.0006139, and for the period from August 2 to August 21, *r*^2^= 0.5932367, where *r* stands for the sample correlation coefficient. Using the Student’s *t* test, with

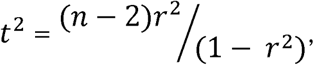

where *n*= 20, we have found that the calculated value of *t* for the first 20 days from June 23 to July 12 comes out as equal to 2.7425 which is greater than the two sided theoretical value of *t* (= 2.101), at 5% probability level of significance for 18 degrees of freedom. Therefore we conclude that during this period the null hypothesis is to be rejected, and that there is a significant linear relationship between Δ*z*(*t*) and *t*. This is equivalent to concluding that the value of the regression coefficient (– 0.00019003) is significantly different from 0.0.

For the next 20 days, from July 13 to August 1, we can see that *t* = 0.1051 which is far smaller than the tabulated value of *t* mentioned above. Therefore we are more than 5% sure that the null hypothesis is true, and therefore we conclude that there is no significant linear relationship between Δ*z*(*t*) and *t*. This is equivalent to concluding that the value of the regression coefficient (– 0.00000889) is not significantly different from 0.0.

For the last 20 days, from August 2 to August 21, it was found that *t* = 5.1236 which is greater than the tabulated value of *t* mentioned above. Therefore we conclude that during this period the null hypothesis is to be rejected, and that there is a significant linear relationship between Δ*z*(*t*) and *t*. Thus we can say that the value of regression coefficient (– 0.0003921) is significantly different from 0.0.

From equations (1), (2) and (3), it was apparent that Δ*z*(*t*) had a decreasing linear trend in the first 20 days, a constancy in the second 20 days and a decreasing linear trend in the third 20 days again, and that the rate of decrease in the third 20 days was about 2 times the rate of decrease in the first 20 days. Constancy of the trend during the second 20 days is clear from the estimated value of the parameter *a* in *z*(*t*)= *log*_*e*_ *N*(*t*) as it has come out to be 0.034550, and the estimate of the parameter *b* was very negligibly small, (= – 0.00000889) in equation (2), while the average of Δ*z*(*t*) during these 20 days was 0.034458 which is almost equal to the estimated value of the parameter *a*.

We have studied the Indian COVID-19 spread data for sixty consecutive days, partitioning the data into three equal parts, with reference to time as the independent variable. However, in the cumulative increase of the number of cases, other than time there actually may be other parameters involved. For example, the spread rate may actually be more among the economically poor people. Therefore the cumulative total number of the cases of this sort of a pandemic is not dependent on time only. We have however studied the situation assuming that time is the only factor affecting the growth of the pandemic.

We have observed that whereas in the first as well as in the last 20 days the negative trend could be clearly seen, in the second 20 days the trend showed constancy which is why the sample correlation coefficient during that period was very nearly zero. This data dependent fact has shown that it is indeed difficult to forecast about the possible time of peaking of the pandemic in India as yet. Indeed, we have already mentioned that we separated the data into three equal parts just to find out this particular uncertainty regarding retardation of the growth of the pandemic in India even by the end of August, 2020.

From equation (3), we can see that for *t* = 78 we get Δ*z*(*t*) = 0. The equations of regression are fitted because they are supposed to be useful in forecasting. Forecasting about the time of peaking of COVID-19 in India may not be very perfect based on such a simple equation of regression. However, the growth cannot continue till Δ*z*(*t*) becomes zero, for that would mean that the total number would suddenly become constant thereafter, which is not possible. Therefore, the change from the exponential to a logarithmic pattern can be expected to take place in India much before 78 days counting from August 2. It should not go beyond that time limit unless something beyond control does take place regarding the spread of the disease. We may perhaps expect that before the end of September, the COVID-19 situation in India should start to retard. In Table-5, we have shown the values of Δ*z*(*t*) for 7 days from August 22 to August 28 to observe whether there is any shift from the trend that has been found from the data in Table-3.

**Table-5:** Values of Δ*z*(*t*) in India from August 22 to August 28, 2020.

| Dates | $N(t)$ | $z(t)$ | $\Delta z(t)$ |
| --- | --- | --- | --- |
| August 28 | 3461240 | 15.05714 | 0.022399 |
| August 27 | 3384575 | 15.03474 | 0.02296 |
| August 26 | 3307749 | 15.01178 | 0.023243 |
| August 25 | 3231754 | 14.98854 | 0.02091 |
| August 24 | 3164881 | 14.96763 | 0.019042 |
| August 23 | 3105185 | 14.94858 | 0.020086 |
| August 22 | 3043436 | 14.9285 | 0.023292 |

From Table-5, the average value of Δ*z*(*t*) for the period from August 22 to August 28, 2020, can be seen to be 0.021705. It can be seen that from the average 0.026449 found during the period from August 2 to August 21, it has come down to 0.021705 during these 7 days. It means that a constancy of the rate of the nearly exponential increase in the total number of COVID-19 cases in India is going to be attained soon, after which the pattern would start taking a nearly logarithmic growth which is commonly known as curve flattening. One point may however be noted in Table-5. The values of Δ*z*(*t*) are not really showing a reducing trend, and if that continues for reasons that can occur due to economic heterogeneity of the population, our prediction might go wrong. We have considered the total number of cases as something dependent on time only, and not on economic and geographical heterogeneity.

In Table-6, we have shown the forecasts of the total number of cases for 10 days in India, taking August 28 as the base date and Δ*z*(*t*)= 0.021705, the average of the values of Δ*z*(*t*) from August 22 to August 28. In Fig. 2, we have shown a comparison of the forecasts with the actual values observed later.

**Table-6:**
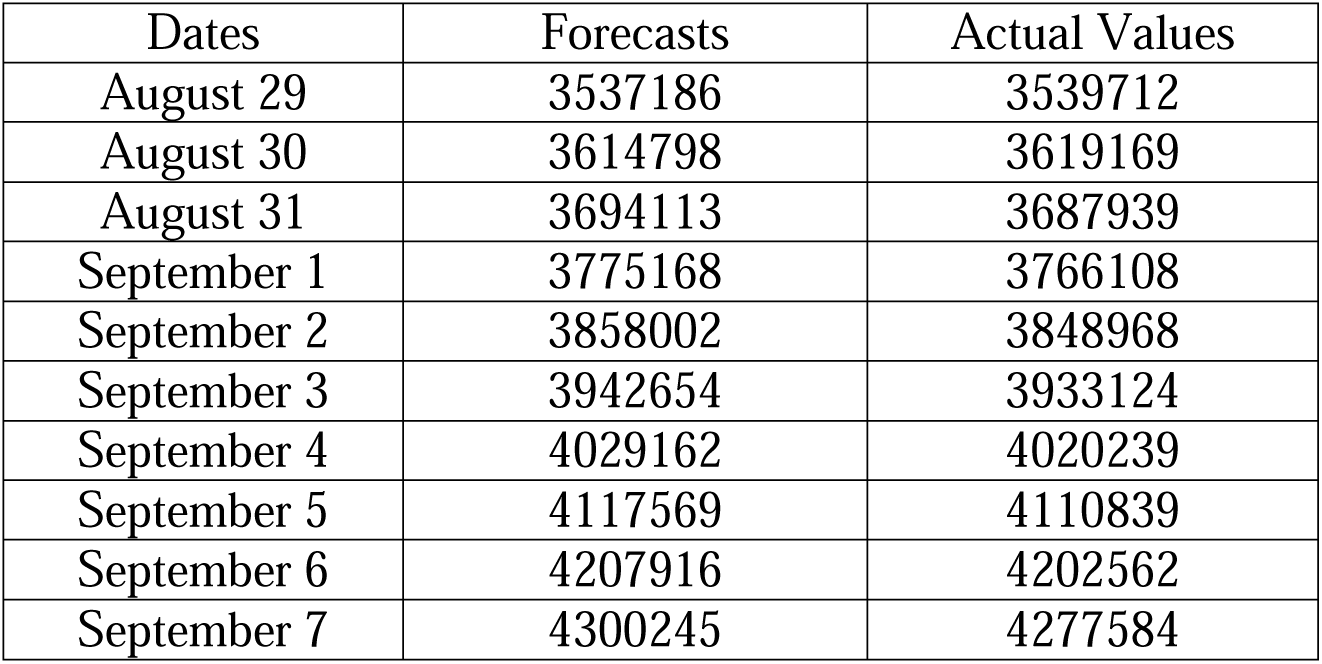
Forecasts for August 29 to September 7

**Fig. 2:**
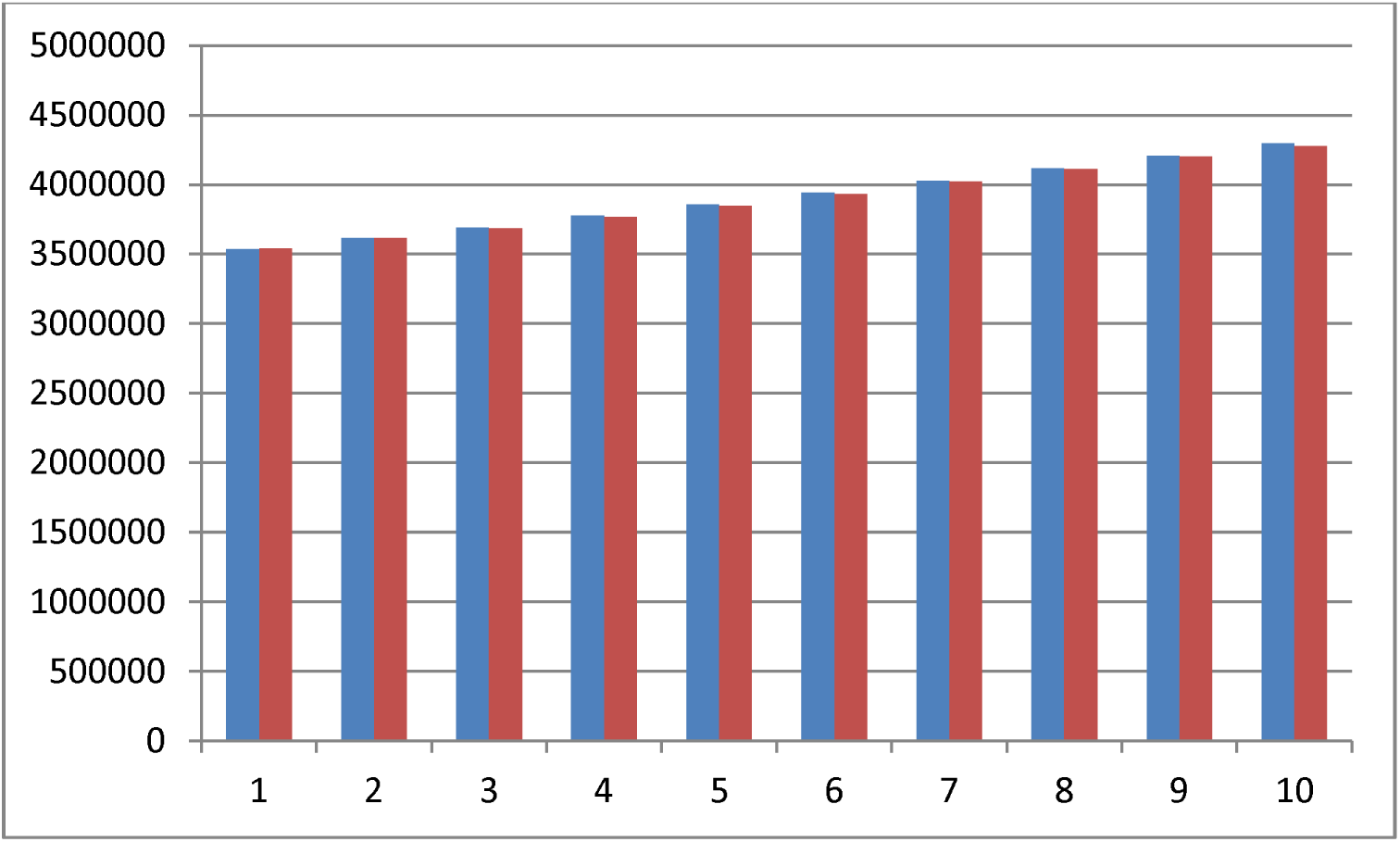
Comparison of the forecasts and the actual values respectively from August 29 to September 7

As we can see from Table-5, the values of Δ*z*(*t*) are actually showing an increasing trend and therefore these forecasts may actually be underestimations of the reality. Indeed even in the 20 days from July 13 to August 1, Δ*z*(*t*) was following a decreasing trend as can be seen from equation (2), although the coefficient of regression of Δ*z*(*t*) on *t* was found statistically insignificant. But during the period from August 22 to August 28, the trend is not a decreasing one. Therefore if the situation continues to remain so, the forecasts would be underestimations only. Accordingly, if the non-decreasing trend of Δ*z*(*t*) continues our forecast regarding the time of peaking also would not work.

If during these 10 days, our forecasts start becoming overestimations, that should be treated as a signal that the change from nearly exponential to nearly logarithmic is perhaps about to start. As can be seen from Fig. 2, the forecasts of the total number of cases from August 29 to August 30 were slight underestimations as expected, and thereafter the forecasts up to September 7 were slight overestimations, and we have mentioned that if these forecasts start overestimating the actual values then that should mean that the change towards improvement of the current situation has perhaps started.

## 4. Conclusions

The first COVID-19 cases were reported in India [4] on February 15, 2020. The spread pattern is however still uncertain in India. We have considered the total number of COVID-19 cases in India for 60 consecutive days, from June 23 to August 21, 2020. We have seen that the first order differences of the natural logarithm of the cumulative total of COVID-19 cases in India have shown statistically significant decreasing linear trend from August 2 to August 21. There may actually be poverty related factors affecting the growth of the total number of cases in India. But if we consider that the growth is dependent on time only, then it can be seen that *b* in *N*(*t*) = exp (*a* + *bt*), *a, b* > 0, *t* ≥ 0, where *N*(*t*) is the total number of cases at time *t*, is decreasing linearly in time. Regression analysis has shown that within less than 78 days starting from August 2, the spread pattern of the pandemic would change from exponential to logarithmic. Indeed, if nothing abnormal happens, the change would occur much before this specified time, because the growth cannot just suddenly be a constant before changing of the pattern from nearly exponential to nearly logarithmic. We have made forecasts of the total number of cases for the duration from August 29 to September 7, but they may be underestimations unless the situation observed from the data from August 22 to August 28 does not change.

## Data Availability

The data were taken from Worldometers.info

